# Conventional vs Virtual Reality in Laparoscopic Training: Study Protocol of a Randomized Controlled Study

**DOI:** 10.64898/2025.12.22.25342766

**Authors:** Sofie Kleiser, Julio César Moreno-Alfonso, Hanna Garnier, Hilmican Ulman, Ophelia Aubert

## Abstract

**Background:** The acquisition of laparoscopic skills is a key component of modern pediatric surgical training, yet structured hands-on exposure remains limited. Conventional box training is widely used but may be constrained by suboptimal visualization and ergonomics. Virtual reality (VR)-based approaches offer immersive visual perspectives and may enhance skill acquisition, motivation, and ergonomic performance. However, robust comparative data on VR-assisted versus conventional laparoscopic training remain limited.

**Methods:** This randomized controlled study evaluates the effectiveness of VR-assisted laparoscopic training compared with conventional training. 40-60 participants, including students, physicians and laypersons without prior laparoscopic experience, will be randomized to either a conventional training group or a VR-assisted training group. Both groups will complete a structured three-week training curriculum using pediatric laparoscopic box trainers and standardized tasks, including PEG transfer, laparoscopic suturing, cyst dissection and a validated esophageal atresia model. The intervention group will use 360 training videos and a VR headset, while the control group will train using standard 2D video material. Pre- and post-training assessments will be video-recorded and evaluated in a blinded manner using validated objective performance metrics (GOALS and OSATS), task-specific error definitions and time to task completion. Secondary outcomes include participant satisfaction, motivation, mental workload and musculoskeletal strain assessed with standardized questionnaires. All data will be pseudonymized and stored securely.

**Conclusion:** This study aims to provide evidence on the role of VR-assisted training in laparoscopic skill acquisition and ergonomics. The findings may inform future training curricula in pediatric and minimally invasive surgery.

## Introduction

The acquisition of laparoscopic skills is a fundamental component of contemporary surgical training, including pediatric surgery. Compared with open procedures, laparoscopy requires distinct psychomotor abilities such as depth perception and instrument handling under indirect visualization ^1,2^. These skills are associated with a prolonged learning curve, particularly for trainees with limited operative exposure ^1,2^. This challenge may be accentuated in pediatric surgery, where caseloads are lower.

Simulation-based training has therefore become an established element of surgical education, enabling structured, repetitive and safe practice independent of clinical case availability. Conventional laparoscopic box training using two-dimensional (2D) video systems is widely implemented, and objective rating instruments such as OSATS and GOALS support standardized assessment of technical performance ^3,4^. Virtual reality (VR)-assisted approaches have been introduced as an adjunct to conventional simulation, aiming to improve spatial orientation ^5-7^. Evidence from adult surgical training shows that VR-based simulator training can translate into improved operative performance in some contexts, but findings are not uniform across studies and vary with design, tasks, and outcome definitions ^8^. In pediatric surgery, data on laparoscopic training remain limited, and few randomized studies combine blinded objective technical assessment with standardized measures of workload and physical strain. To address this gap, the present randomized controlled study employs validated technical performance scales in combination with established measures of mental workload and musculoskeletal strain in the context of pediatric surgery ^3,4,9,10^.

## Methods

Participants will be recruited starting 2025 and attend the training course until the projected sample size has been achieved in participating pediatric surgical departments at the University Medical Center Mannheim, the University Hospital of Navarra, the Medical University of Gdansk and the Ege University Faculty of Medicine. The study was approved by each respective ethics committee and the study was registered in the German Trial Register.

### Participants

Based on a power analysis using comparable training studies, a number of approximately 40-60 participants is needed . Participation in the study is voluntarily and participants must give their written consent. Block randomization will be done using the online generator “Research Randomizer” (https://www.randomizer.org/).

<u>Inclusion criteria:</u> students, physicians, or laypersons no prior laparoscopic experience willingness to complete the training curriculum.

<u>Exclusion criteria:</u> attendance at a course in minimally invasive surgery involvement in study planning.

### Study Design

#### Control group

Laparoscopic training will be conducted using the pediatric LaparoscopyBoxx system in combination with a smartphone-based video camera ^11^. Training videos are provided in a 2D format, with a smartphone serving as the display screen during practice sessions. Participants have continuous access to the training materials throughout the study period. Pre- and post-training assessments are recorded using the same device employed as the display screen or a compatible interconnected device, all in a 2D viewing format.

#### Intervention group

Laparoscopic training will be performed using the pediatric LaparoscopyBoxx system, together with a 360° video camera (Insta360 One X2) and a VR headset (Meta Quest 3). Participants will view a 360° training video via the Meta Quest 3 to provide a three-dimensional impression of the training environment. During hands-on practice, the VR headset will be used as the display while an interconnected compatible device provides the camera feed, and participants will have continuous access to the training materials throughout the study period. Pre- and post-training assessments will be conducted using the Meta Quest 3 headset; recordings will be captured using the smartphone serving as the camera.

Before the start of the first training, the participant will fill out a questionnaire to assess surgical skills and motivation, demographical and lifestyle data (sleeping habits, sports, musical instrument practice, gaming, average caffeine & nicotine consume) and prior laparoscopic training experience. The training program extends over 3 weeks. The following tasks will be practiced: PEG transfer, laparoscopic suturing, cyst dissection and a validated laparoscopic model of esophageal atresia. The correct execution of each task will be demonstrated in a video before the pre-test and can be rewatched and paused at any time if needed. During training, the video material will also be available for the participant.

### Tasks

#### PEG transfer

For the simulated laparoscopic transfer task, the participant needs to move a rubber ring using two instruments. He will start by picking up the ring on the right side of the board, then passing it to the instrument in the other hand, placing it on the left side of the board. After transferring the ring, the process will be repeated, this time transporting the ring from the outside to the inside. If the ring is dropped within the field of view, it must be picked up using the same instrument that dropped it. The timing will begin with the first touch of the ring and will end on its correct placement.

#### Laparoscopic suturing

The membrane is attached on all four sides. Using a needle holder and a needle with suture, an intracorporeal single-button stitching must be placed at the marked point by placing a single knot f irst and two opposing knots following and cut the end. The task is accomplished once the participant performed a complete stitch.

#### Cyst Dissection

Two standard plastic balloons are used to represent tissue layers. The inner balloon is placed inside the outer balloon and inflated slightly with air. Using laparoscopic instruments, the participant carefully incises the outer balloon to allow for controlled peeling while preserving the integrity of the inner balloon. The task is completed once the inner balloon is fully exposed without damage.

#### Esophageal atresia (EA) model

For the EA model, two balloons will be used ^12^. The small end side from one balloon (±5 mm from the edge) and the wide end side from the other (also ±5 mm from the edge) will be cut off. Both balloons will be attached to the suturing pads with a gap of 2–3 mm between the balloon ends. The task is to open the pouch and place 3 sutures for the anastomosis.

PEG transfer and laparoscopic suturing should be practiced at the beginning to practice the handling of the instruments. Cyst dissection and esophageal atresia are to be trained later.

Over the course of three weeks, the participant will receive 6 training sessions. In the first session the participant must answer the questionnaire to assess the baseline characteristics as described above and undergo the pre-training test in which the PEG transfer, laparoscopic suturing and cyst dissection are performed once. In the following sessions the participant can train the various units as described above. The last session will end with the post training test and answering of the follow-up questionnaires to assess the outcome.

### Outcomes

The performance will be analyzed according to the following criteria: the overall time to task completion, the time to task completion of each task and the errors made per task.

Errors / success are defined for each task as follows:

- Peg transfer: number of times PEG dropped
- Laparoscopic suturing: knot tightness (tight knot - knot loose under tension - visibly loose knot)
- Cyst dissection: Removal of the outer layer (inner ballon damaged – insufficient removal of the outer balloon - complete removal of the outer balloon)
- Esophageal atresia model: stitches (all correct - one incorrect - two incorrect - all three stitches incorrect) knot tightness (tight knots - knots loose under tension - visibly loose knots) adaption of the endings (completely adapted - partly adapted - not adapted/curved upwards)

Furthermore, the outcome will be evaluated with two quality assessment tests. The Global Operative Assessment of Laparoscopic Skills (GOALS) measures 5 items (depth perception, ambidexterity, efficiency, tissue handling, autonomy) using a Likert scale (1-5) ^3^. The maximum score to be achieved is 25 and corresponds to an excellent surgical performance. The minimum score is 5 and corresponds to poor surgical performance. This test will be used to analyze the video recordings. The analysis will be done in a blinded way. In the Objective structured assessment of technical skill (OSATS) a total of 7 items are examined, which includes respect of tissue, time and movement, handling of instruments, knowledge of the instruments, use of the assistant, flow and procedure of the operation and advance planning and knowledge about the specific operation ^4^. The assessment is carried out using a Likert scale (1-5). The maximum score is 35 points and corresponds to an excellent result, while the minimum score is 7 and corresponds to poor operational performance.

Participant satisfaction (accuracy and usefulness of the training model, enjoyment of the training, ergonomics of the training) is assessed using the Questionnaire on Current Motivation ^13^. This questionnaire consists of 18 items to measure four motivational factors: anxiety, probability of success, interest, and challenge using a 5- point Likert scale and a standardized questionnaire.

Additionally, a questionnaire was developed to evaluate the participants’ experiences with the two training methods, which assesses with 12 questions the well-being during the exercises, the usefulness of the video material and instructions, the perceived effects and if the participants would recommend their training method to others. The questionnaire consists of multiple-choice items with a five-point-Likert scale, open-ended questions and questions with binary responses that provide both quantitative and qualitative insights into user satisfaction and the effectiveness of the training.

The mental load and musculoskeletal tension will be recorded using the standardized assessment of the Borg Musculoskeletal Scale and the NASA task load index (NASA-TLX) ^9,10^. The NASA-TLX is a questionnaire to assess mental, physical, temporal demands, performance, effort and frustration levels.

## Discussion

This randomized controlled study is designed to compare VR-assisted and conventional laparoscopic training within a structured training curriculum. We anticipate that VR-assisted training may result in differences in selected performance domains, particularly those related to spatial orientation, efficiency and ergonomics. Improvements may be reflected in objective performance metrics such as task completion time, error rates and validated global rating scales including GOALS and OSATS ^3,4^.

Beyond technical performance, VR-assisted training may influence subjective outcomes, including perceived workload, physical strain and user experience. Enhanced visual immersion and flexible viewing conditions could potentially reduce mental workload and musculoskeletal discomfort during training, as assessed by standardized instruments such as the NASA-TLX and the Borg Musculoskeletal Scale ^9,10^. At the same time, it is acknowledged that the use of VR headsets may introduce novel sources of discomfort or fatigue for some participants, which will be captured through post-training questionnaires. The inclusion of blinded video-based assessment and task-specific error definitions is intended to minimize assessment bias and enhance methodological robustness.

Several limitations should be considered. The study population includes participants with heterogeneous backgrounds and results may not be directly generalizable to surgical residents or experienced surgeons. In addition, while standardized tasks allow objective comparison, they do not fully capture the complexity of real operative scenarios. Nevertheless, by integrating objective performance measures with validated assessments of workload, strain and user experience, this study aims to provide a comprehensive evaluation of VR-assisted versus conventional laparoscopic training.

Overall, the anticipated results are expected to contribute to understanding of the potential role of VR-assisted simulation within laparoscopic training curricula.

## Conclusion

This randomized controlled study will compare VR-assisted and conventional 2D laparoscopic training using validated performance assessments and standardized measures of workload and user experience. The results are expected to inform evidence-based integration of VR into laparoscopic training curricula, including pediatric surgical education.

## Data Availability

All data produced in the present study are available upon reasonable request to the authors.

## Literature

1. Vajsbaher T, Schultheis H, Janssen S, et al. The development of visuospatial abilities and their impact on laparoscopic skill acquisition: a clinical longitudinal study. Surgical endoscopy. Dec 2022;36(12):8908–8917. doi:10.1007/s00464-022-09328-1

2. Beattie KL, Hill A, Horswill MS, Grove PM, Stevenson ARL. Aptitude and attitude: predictors of performance during and after basic laparoscopic skills training. Surgical endoscopy. May 2022;36(5):3467–3479. doi:10.1007/s00464-021-08668-8

3. Vassiliou MC, Feldman LS, Andrew CG, et al. A global assessment tool for evaluation of intraoperative laparoscopic skills. Am J Surg. Jul 2005;190(1):107–13. doi:10.1016/j.amjsurg.2005.04.004

4. Martin JA, Regehr G, Reznick R, et al. Objective structured assessment of technical skill (OSATS) for surgical residents. Br J Surg. Feb 1997;84(2):273–8. doi:10.1046/j.1365-2168.1997.02502.x

5. Lungu AJ, Swinkels W, Claesen L, Tu P, Egger J, Chen X. A review on the applications of virtual reality, augmented reality and mixed reality in surgical simulation: an extension to different kinds of surgery. Expert Rev Med Devices. Jan 2021;18(1):47–62. doi:10.1080/17434440.2021.1860750

6. Li T, Yan J, Gao X, et al. Using Virtual Reality to Enhance Surgical Skills and Engagement in Orthopedic Education: Systematic Review and Meta-Analysis. J Med Internet Res. May 30 2025;27:e70266. doi:10.2196/70266

7. Sommer GM, Broschewitz J, Huppert S, et al. The role of virtual reality simulation in surgical training in the light of COVID-19 pandemic: Visual spatial ability as a predictor for improved surgical performance: a randomized trial. Medicine (Baltimore). Dec 17 2021;100(50):e27844. doi:10.1097/md.0000000000027844

8. Larsen CR, Soerensen JL, Grantcharov TP, et al. Effect of virtual reality training on laparoscopic surgery: randomised controlled trial. Bmj. May 14 2009;338:b1802. doi:10.1136/bmj.b1802

9. Borg G. Borg’s perceived exertion and pain scales. Borg’s perceived exertion and pain scales. Human Kinetics; 1998:viii, 104-viii, 104.

10. Hart SG, Staveland LE. Development of NASA-TLX (Task Load Index): Results of empirical and theoretical research. Human mental workload. North-Holland; 1988:139–183. Advances in psychology, 52.

11. Joosten M, Hillemans V, van Capelleveen M, et al. The effect of continuous at-home training of minimally invasive surgical skills on skill retention. Surgical endoscopy. Nov 2022;36(11):8307–8315. doi:10.1007/s00464-022-09277-9

12. Bökkerink GMJ, Joosten M, Leijte E, Lindeboom MYA, de Blaauw I, Botden SMBI. Validation of low-cost models for minimal invasive surgery training of congenital diaphragmatic hernia and esophageal atresia. Journal of Pediatric Surgery. 2020/06/13/ 2020;doi:10.1016/j.jpedsurg.2020.05.045

13. Rheinberg F, Vollmeyer R, Burns B. FAM. Ein Fragebogen zur Erfassung aktueller Motivation in Lern-und Leistungssituationen [Verfahrensdokumentation aus PSYNDEX Tests-Nr. 9004322 und Fragebogen]. Leibniz-Zentrum für Psychologische Information und Dokumentation (ZPID)(eds) Testarchiv Trier: ZPID. 2019;

